# Feasibility of linking markers of dementia-related health in primary care medical records to cognitive function assessed in a specialist dementia service

**DOI:** 10.1101/2022.10.11.22279756

**Authors:** Michelle Marshall, Paul Campbell, James Bailey, Carolyn A Chew-Graham, Peter Croft, Martin Frisher, Richard Hayward, Rashi Negi, Trishna Rathod-Mistry, Swaran Singh, Louise Robinson, Athula Sumathipala, Nwe Thein, Kate Walters, Scott Weich, Kelvin P Jordan

## Abstract

**Objectives:** To assess the feasibility of linking and comparing markers of dementia-related health recorded in primary care electronic health records (EHR) to assessments of cognitive function undertaken in a specialist dementia service.

**Methods:** One thousand patients in a UK secondary care specialist dementia service were invited to take part. Primary care EHR were requested from 72 general practices of consenting patients. Sixty-three previously established individual markers within 13 broader domains of dementia-related health were then extracted from primary care EHR and compared to cognitive assessments scores recorded in the dementia service EHR.

**Results:** 258 (26%) patients consented to take part. At least one cognitive assessment score was recorded for 242 (94%) patients, but primary and secondary care EHR records could only be linked in 93 patients. 56 of these 93 patients had two cognitive assessments scores at least 12 months apart. In the patients with data available for analysis individuals with a higher number of markers and domains recorded in their primary care records had lower mean cognitive assessment scores (range 1.6-2.1 points), and after adjustment for earlier cognitive scores (range 2.0-2.5 points), indicating poorer cognitive function, although differences were not statistically significant.

**Conclusion:** This feasibility study highlights the challenges in obtaining consent and linking primary and secondary care EHR in dementia, and in extracting cognitive function scores from dementia service EHR.

## Introduction

The number of people with dementia is increasing as the population ages and dementia has a large impact on the lives of individuals with the condition as well as their families and caregivers.^1,2^ Strategies have been proposed to prolong independence, reduce hospital admissions, delay nursing home admissions, and prevent early mortality for people with dementia.^3-6^ Information on the course of dementia prior to these long-term outcomes could improve prognosis at an individual patient level, aid planning and monitoring of care for dementia, and allow evaluation of earlier outcomes in research studies including clinical trials in dementia.^1,7^

In many countries primary care is the first point of contact and location of management of common health conditions including dementia. Primary care can play a key role in addressing strategies to improve outcomes for dementia. One potential resource for monitoring the course of dementia in primary care are Electronic Health Records (EHR). Primary care EHR contain information that is routinely recorded in patient encounters. This typically includes coded reasons for consultations, prescriptions, referrals, investigations and tests. In the UK, over 95% of the population are registered with a general practitioner (GP) and the place where most routine chronic disease management including dementia occurs, and so these records are a useful resource for studying how illnesses progress. However, to date, primary care EHR have not been used to research the course of dementia after diagnosis. There is evidence that this might be possible as key comorbidities and signs and symptoms likely to be recorded in primary care have been associated with dementia and could be indicative of disease progression and severity (e.g. malnutrition, fall trauma, neuropsychiatric disorders, sleep disorders.^8,9^ These signs and symptoms are likely to occur prior to more recognised long-term outcomes such as hospital or care home admission, and earlier mortality. Therefore, there is the potential for primary care EHR to be a source of population-wide data on course and prognosis of dementia for research and monitoring and for targeted anticipatory care of individuals.

We have previously established a set of potential primary care EHR markers (categorised into different domains) of dementia progression,^10^ and shown that these are associated with future outcomes such as mortality and hospital admission.^11^ In particular, we found that the number of different domains accumulated in the primary care records in a 12-month period was associated with the occurrence of these future outcomes.^11^ However, an important gap in developing these EHR markers as the basis for epidemiological and intervention studies is to establish their construct validity as markers of actual dementia severity and progression.

In order to address this gap, we have undertaken a feasibility study obtaining and linking primary and secondary care EHR in patients sampled from a secondary care setting where objective assessments of cognitive function had been performed and recorded as part of clinical care. In patients consenting to accessing and linking their records, we compared the results of these assessments with data extracted independently from the primary care EHR of the individuals in this sample. We also assessed the challenges of performing this type of study.

## Materials & methods

### Study population

The CoMed study recruited patients from a secondary care dementia service within South Staffordshire and Shropshire, UK, delivered by the Midlands Partnership NHS Foundation Trust. Written informed consent was obtained from patients with dementia (or personal consultee’s advice for those not able to give consent) to access and link their secondary care dementia service and primary care medical records for research purposes. Ethical approval was obtained by the UK National Research Ethics Service, Wales 7 Committee (REC reference: 18/WA/0423).

Eligible participants met the following selection criteria. Inclusion criteria:

- Aged 18 years and over
- Confirmed diagnosis of dementia recorded in the dementia service medical records
- Assessment by the dementia service in the previous 12 months
- Living in the UK regions covered by three local Clinical Commissioning Groups (CCGs)

Exclusion criteria:

- Lists of potentially eligible patients were screened by clinical care teams to exclude those where contact would likely cause undue distress or harm e.g. palliative care or significant life event
- A recorded indication in their dementia service medical records that they did not wish to take part in research.

One thousand eligible patients were randomly selected and mailed a study information pack by post inviting them to take part in the study, i.e. consent to access and linkage of their primary care and dementia service records, with a reminder sent after two weeks if no response.

### Data collection from medical records

In those consenting to take part in the study, cognitive assessment scores in the 10 years prior to the date of consent were retrieved from the electronic dementia service medical records. Cognitive assessments used by the dementia service included the Mini Mental State Examination (MMSE), Addenbrooke’s Cognitive Examination - III (ACE-III) and Mini Addenbrooke’s Cognitive Examination (MACE).^12-14^ Higher scores for each test reflect better cognitive function. ACE III (range 0-100) and MACE scores (0-30) were converted into standardised MMSE scores (0-30) using previously established conversion methods.^15,16^

Primary care EHR were requested from each consenting patient’s general practice for the 10 years prior to the date of consent was provided. This included all recorded electronic Read codes (a hierarchical coding system used in UK primary care for recording morbidity and processes of care) and prescriptions. EHR were requested in the form of an electronic download at the general practice and transferred to the researchers via NHS email.

### Markers of dementia progression

A list of potential primary care markers (Read coded and prescribed medication) of dementia-related health nested into domains has been established previously. Full methodology is detailed elsewhere;^10^ but included systematic literature searches, consensus exercises including GPs, psychiatrists, epidemiologists and EHR researchers, and analysis of a regional primary care EHR database. Sixty-three potential markers of dementia-related health were grouped into 13 domains (Supplementary Table 1): Care, Home Pressures, Severe Neuropsychiatric, Neuropsychiatric, Cognitive Function, Daily Functioning, Safety, Comorbidity, Symptoms, Diet/Nutrition, Imaging, Increased Multimorbidity (based on polypharmacy), and Change in Dementia-Related Drug.

### Analysis

In consenting patients, the number and proportion of patients with linked primary and secondary care EHR was determined. Then i) the number of patients with at least one cognitive assessment score recorded was established for the cross-sectional analysis and ii) the number of patients with two cognitive assessments scores at least 12 months apart was established for the longitudinal analysis.

We compared the results of cognitive assessments undertaken as part of clinical care in the secondary care dementia service with data extracted independently from the primary care EHR.

For the cross-sectional analyses all consenting participants who had at least one assessment recorded in the dementia service were included. Domains and markers were identified in their primary care EHR for the 12 months before each patient’s most recent cognitive assessment (the “end” score). Patients were then grouped based on the tertile number of domains and of markers recorded in the primary care EHR over that 12-month period. The mean standardised MMSE score for each group and mean differences in scores between groups were calculated using the most recent cognitive assessment score in the dementia service medical records. The relationships between cognitive assessment scores and recording of individual domains were also determined.

For the longitudinal analyses the sub-group of consenting participants who had at least two assessments recorded in the dementia service at least 12 months apart were analysed. Records of domains and markers were identified in primary care records between the dates of a patient’s earliest (start) and most recent (end) cognitive assessment at the dementia service. Patients were again grouped based on the tertile number of domains and of markers recorded in the primary care EHR over that period. Mean standardised MMSE end score and mean differences in scores between groups (with 95% confidence intervals) were derived adjusting for the earliest recorded score (the “start” score) using analysis of covariance. Finally, the relationships between most recent cognitive assessment score and recording of individual domains, adjusted for earliest cognitive assessment score, were also determined.

## Results

Of the 1000 patients invited to take part, 258 (26%) consented (Figure 1). Two-hundred and forty-two (94%) patients had one or more cognitive assessment scores recorded in their dementia service medical records. Primary care EHR were obtained and linked to dementia service medical records for 93 (38%) of these 242 patients from 34/72 (47%) GP practices and they formed the main sample in which the cross-sectional analysis was undertaken. There was no response from 30 (42%) practices covering 121 (50%) patients. There were 8 (11%) GP practices that did make contact but who did not contribute EHR data. The main reasons for this were being too busy (4 GP practices; 12 patients), incompatible systems for electronic download (2 GP practice; 8 patients) and inability/against practice protocol to send electronic data (2 GP practices; 8 patients). Age and gender distributions were comparable between those with and without primary care EHR information, but the diagnosis duration to MMSE end score was shorter and the end (most recent) median MMSE score was slightly higher indicating better cognitive function in those with linked primary care information (Table 1).

**Table 1.**
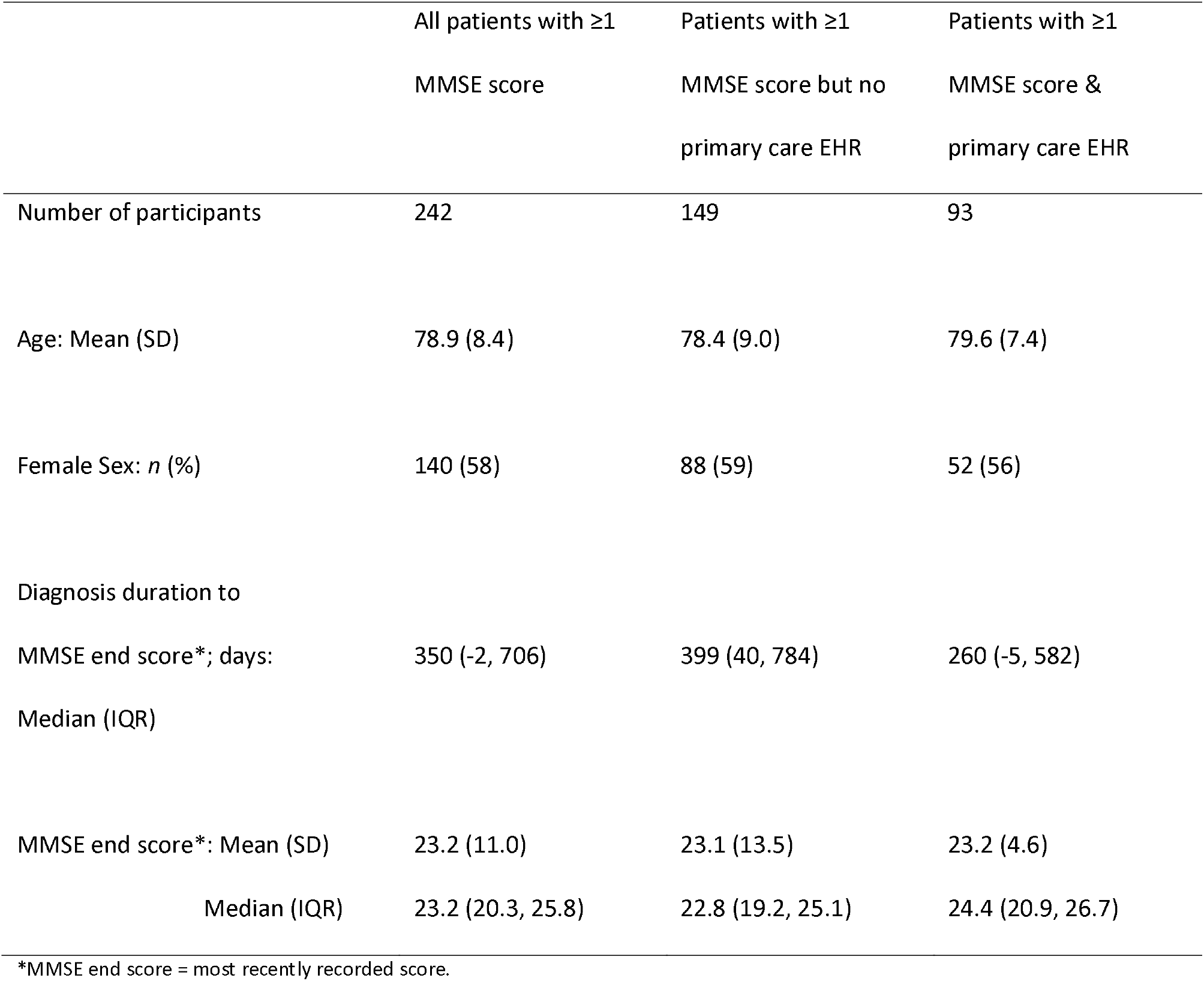
Descriptive characteristics of study participants overall, and in those with and without linked primary care EHR.

**Figure 1.**
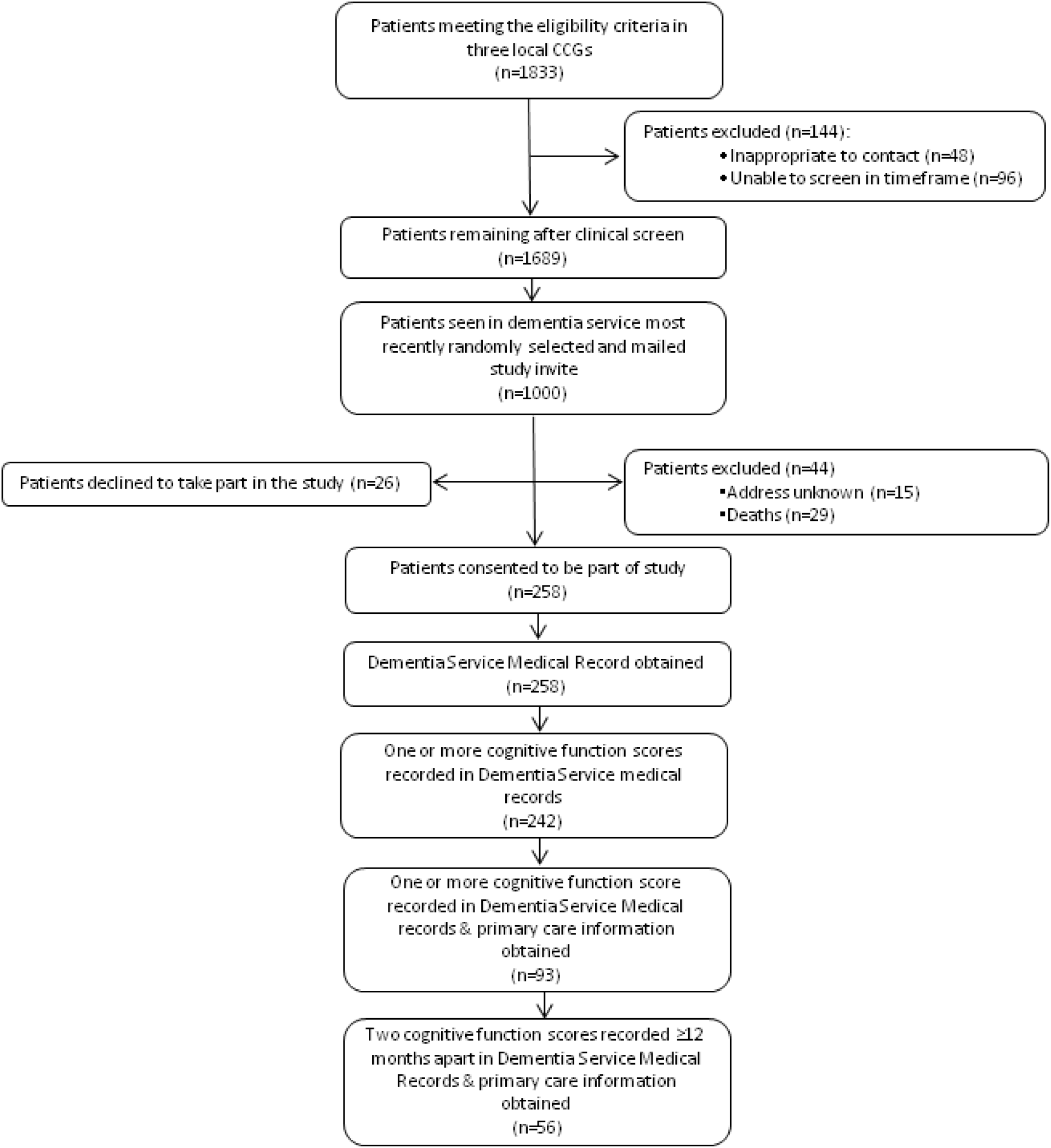
Study flowchart of recruitment.

In the cross-sectional analysis, individuals with the highest numbers of markers (≥5) and domains (≥4) recorded in their primary care records in the 12 months before their most recent dementia service assessment had lower mean MMSE end scores by 2.1 (markers) and 1.6 (domains) points, respectively, than those with the fewest (0-2 markers or domains) indicating poorer cognitive function (Table 2). However, differences were not statistically significant.

**Table 2.** Relationship between MMSE score and the number of domains and markers (n=93)

|  | Number | n | Median MMSE<br>end score (IQR) | Mean MMSE<br>end score (SD) | Mean difference in<br>MMSE end score<br>(95% CI) |
| --- | --- | --- | --- | --- | --- |
| Domains<sup>a</sup> | 0-2 | 37 | 24.4 (21.8, 26.2) | 23.6 (3.6) | Ref |
|  | 3 | 25 | 26.2 (22.1, 27.8) | 24.0 (4.8) | 0.4 (-1.9, 2.8) |
|  | ≥4 | 31 | 23.8 (18.0, 25.6) | 22.0 (4.6) | -1.6 (-3.8, 0.6) |
| Markers<sup>a</sup> | 0-2 | 29 | 24.4 (21.2, 26.4) | 23.6 (3.9) | Ref |
|  | 3-4 | 38 | 25.1 (21.9, 27.2) | 24.0 (4.2) | 0.4 (-1.8, 2.6) |
|  | ≥5 | 26 | 23.5 (18.0, 25.4) | 21.6 (4.6) | -2.1 (-4.5, 0.3) |

<sup>a</sup> Recorded in the 12m before most recent MMSE assessment date. MMSE end score = most recently recorded score.

Fifty-six patients had two cognitive assessments scores recorded in the dementia service medical records a minimum of 12 months apart and had primary care information obtained. These patients formed the sub-sample in which the longitudinal analysis was undertaken. Median time between start and end assessments was 783 (IQR 555, 1116) days. Mean differences in most recent MMSE scores after adjustment for earliest MMSE score, comparing those with the most recorded markers (≥7) and domains (≥6) to those with the fewest, were 2.0 and 2.5 points, respectively (Table 3). This suggest more cognitive function decline in those with more recorded markers and domains, however the differences were not statistically significant.

**Table 3.** Relationship between MMSE score over time and the number of domains and markers (n=56)

|  | Number | n | Mean | Unadjusted mean | Adjusted mean |
| --- | --- | --- | --- | --- | --- |
|  |  | patients | MMSE end | difference in MMSE | difference in MMSE |
|  |  |  | score | end score | end score<sup>a</sup> |
|  |  |  |  | (95% CI) | (95% CI) |
| Domains<sup>b</sup> | 1-3 | 20 | 24.5 | Ref | Ref |
|  | 4-5 | 26 | 23.0 | -1.6 (-4.5, 1.3) | -1.0 (-3.4, 1.3) |
|  | ≥6 | 10 | 21.1 | -3.5 (-7.3, 0.3) | -2.5 (-5.5, 0.6) |
| Markers<sup>b</sup> | 1-4 | 22 | 24.2 | Ref | Ref |
|  | 5-6 | 16 | 23.4 | -0.8 (-4.1, 2.4) | -0.4 (-3.0, 2.2) |
|  | ≥7 | 18 | 21.8 | -2.4 (-5.5, 0.8) | -2.0 (-4.5, 0.5) |

<sup>a</sup> Adjusted for MMSE start (earliest) score; <sup>b</sup> Recorded between dates of start and end score. End score = most recently recorded score.

Individuals in the cross-sectional sample (n=93) who had markers recorded in the domains of Daily Functioning, Safety, Care, and Diet/Nutrition in the 12 months before their dementia service assessment had lower mean MMSE end scores by 3.5 to 7.6 points, indicating poorer cognitive function compared to individuals that did not have markers recorded from these domains (Table 4). However, the number of people recorded with these domains was low.

**Table 4.** Relationship between MMSE score and individual domains (n=93)

|  | n patients | Mean MMSE end score |  | Mean difference (95% CI) |
| --- | --- | --- | --- | --- |
| Domain<sup>a</sup> | with |  |  |  |
|  | recorded | Domain | Domain |  |
|  | domain | absent | present |  |
| Care | 13 | 23.7 | 20.2 | -3.5 (-6.2, 0.9) |
| Home Pressures | 0 | 23.2 | c | c |
| Severe Neuropsychiatric | 2 | 23.3 | c | c |
| Neuropsychiatric | 42 | 23.1 | 23.3 | 0.3 (-1.6, 2.2) |
| Cognitive Function | 35 | 22.4 | 24.5 | 2.1 (0.2, 4.0) |
| Daily Functioning | 5 | 23.6 | 16.1 | -7.6 (-11.4, -3.7) |
| Safety | 7 | 23.5 | 19.0 | -4.6 (-8.0, -1.1) |
| Comorbidity | 54 | 22.6 | 23.6 | 1.0 (-0.9, 2.9) |
| Symptoms | 26 | 23.3 | 23.0 | -0.3 (-2.4, 1.8) |
| Diet/Nutrition | 17 | 23.9 | 20.2 | -3.7 (-6.0, -1.3) |
| Imaging | 17 | 23.2 | 23.0 | -0.2 (-2.7, 2.2) |
| Increased Multimorbidity<sup>b</sup> | 40 | 23.5 | 22.8 | -0.6 (-2.5, 1.3) |
| Change in Dementia-related Drug<sup>b</sup> | 21 | 23.1 | 23.6 | 0.6 (-1.7, 2.8) |

<sup>a</sup> Recorded in 12 months before most recent MMSE assessment date; <sup>b</sup> Compared to previous 12 months; <sup>c</sup> Not presented
as prevalence was less than 5 people. MMSE end score = most recently recorded score.

In the longitudinal analysis (n=56), reduced mean scores on the most recent MMSE assessment persisted for patients with recorded markers in the domains of Daily Functioning, Safety, Care, and Diet/Nutrition after adjustment for earliest recorded MMSE score by 1.7 to 3.3 points, showing they had more cognitive function decline compared to individuals that did not have markers in these domains (Table 5). However, the number of patients with these domains were again low and differences were only statistically significant for the Safety domain.

**Table 5.**
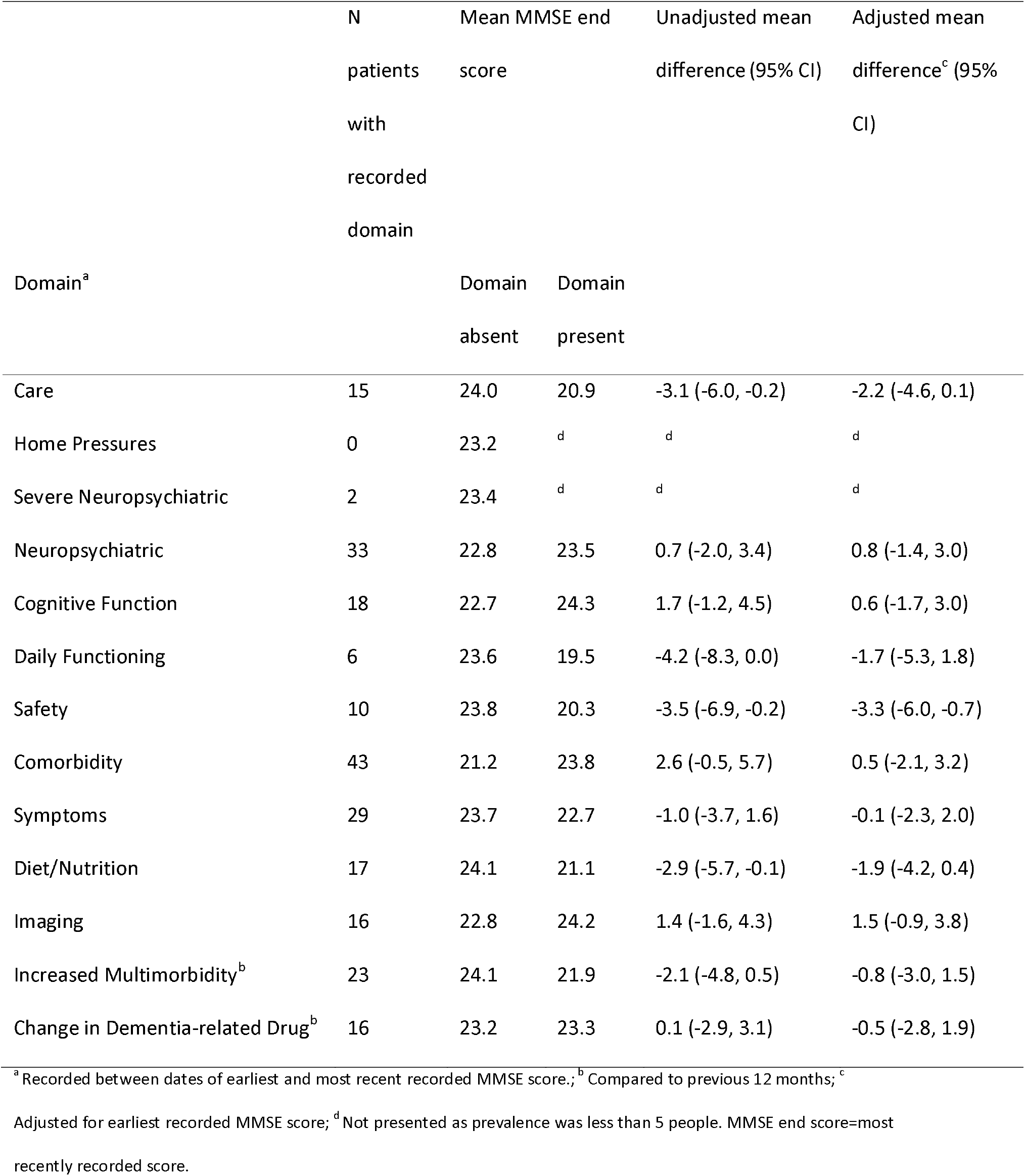
Relationship between MMSE score over time and individual domains (n=56)

## Discussion

This study aimed to pilot linking and comparing potential markers of dementia progression routinely recorded in primary care against cognitive assessments undertaken in a specialist dementia service. There were difficulties in obtaining primary care information which meant that linked primary and secondary care EHR could only be obtained in just over a third of consenting patients. Further to this, fewer than expected patients had repeated cognitive assessment scores that were at least 12 months apart recorded in the dementia service medical records. While the study was underpowered, those with a higher number of domains and markers recorded in primary care had trends towards poorer cognitive function as assessed in the dementia service which suggests the domains and markers are associated with greater disease progression. These differences were not statistically significant, but the findings do concord with our previous validation study which showed that the number of recorded domains early after diagnosis were strongly associated with long term outcomes of hospital admission, palliative care and mortality.^11^

This study used information routine collected as part of primary care to investigate a rigorously developed set of domains and markers. This approach reduced the burden on the patient with dementia and their caregiver who were asked only for consent to access and link medical records. Previous dementia studies have recruited by post with response rates from UK and other countries ranging between 22.3 (UK) and 31.3% (Norway);^17,18^ this method was selected as it was felt it would be less time consuming than recruiting through dementia service clinics and would not add extra burden to clinicians or patients in the consultations in which there are already often time pressures. While our response rate of 25.2% is generally in line with the previous dementia studies recruiting by post, and comparable to other large surveys, the response to our study was low.^17-19^ Information including study packs handed out by dementia service staff at clinical appointments could potentially increase response. A study population of 258 could have given us sufficient power to undertake the planned analysis however, there was difficulty in obtaining information from general practices. This was despite providing instructions and technical support for undertaking the electronic download of GP record data. This is likely to reflect time pressures but may also relate to lack of experience with research and concern over releasing records in more research-naïve practices. This reduced the power of our study, and particularly limited our assessment of individual domains.

In this study there were also a number of challenges related to the lack of standardisation and completion of cognitive assessment measures utilised within the secondary care dementia service.^4^ Firstly, mapping ACE-III and MACE to the MMSE was possible based on published formulae, however this method has not been extensively validated and some patients had values outside the range of the MMSE once standardised. Secondly, many patients had only one (or no) cognitive assessment score and this may reflect that in clinical practice it is the individual areas covered by the measures that are more important (i.e. the indicators of specific components of memory function), rather than the full scores themselves. Added to that is the potential for patients to be unwilling or not able to complete the full test. This study has also focussed on those with diagnosed dementia. While advances have been made to improve the timely detection of dementia within primary care;^20^ a significant proportion (∼ 40%) will be undetected and undiagnosed, and those with more severe dementia may be more likely to be diagnosed.^21,22^

Despite the reduced sample size, the domains of Care, Daily Functioning, Safety, and Diet/Nutrition showed trends of poorer cognitive assessment scores in those with markers from these domains recorded in primary care. The mean differences seen in MMSE scores between individuals with and without markers recorded in these domains were clinically significant when compared to recommended levels of important differences (between 1 and 3 MMSE points).^23^ Importantly, these domains were also found to be associated with longer term outcomes of hospital admission, palliative care and mortality in a previous validation study in a UK primary care database.^11^ However, there was a low number of people with these domains recorded and so this current study should be viewed as exploratory with its findings requiring confirmation in larger studies.

Our findings, in this and our previous study, suggest particular domains and markers impact on outcomes for those with dementia.^11^ This has also been found in previous non-EHR research internationally. For example, changes in care including shared decision making and advanced care planning have been found to be associated with care home admission and palliative care,^24,25^ and caregiver coping and stress are associated with mortality in the person with dementia.^26^ Common markers in the Safety domain are falls and fractures and are likely to reflect the increased vulnerability in this population. Falls have previously been associated with increased rates of hospitalisation and mortality.^27-29^ Nutrition has previously been associated with the longer-term outcome of mortality in people living with dementia.^30,31^ This supports the creditability that these domains may be valid indicators of poorer short-term and long-term outcomes in dementia.

A potentially unexpected finding was that individuals who had one or more primary care markers recorded in the cognitive function domain had higher (better) MMSE scores in the cross-sectional analysis, although this was not apparent when adjusted for earlier MMSE score in the longitudinal analysis. It is possible cognitive function markers are recorded in primary care more commonly in the earlier stages of the disease as part of the diagnosis process.

The challenges in gaining consent and collating EHR information in this study can inform similar future research, including the larger studies needed to confirm the findings of this study. Improved collection of primary care information may be possible by reversing the approach taken here and determining the initial study population at general practices, who agree a priori to release records of consenting patients and are located within the dementia service’s catchment area, rather than at the dementia service. This would mean gaining agreement from a large number of practices and hence be more resource intensive. It may also help to target GP practices that have previously been involved in research as they will have greater understanding and appreciation of being involved in research, and potentially be more supportive. In the future, improved integration of records between primary and secondary care may also help studies like this. However, future research studies may also need to consider patient-reported data collected by either survey or interview and linked to EHR in order to explore in-depth the association between dementia assessments, patient and carer information, and EHR data.

Despite the challenges, the findings from this study of linked primary care and secondary care dementia service medical records and our previous study using a national EHR database suggest these markers and domains recorded in primary care do reflect disease progression in dementia. Further research is needed to assess how these markers and domains may be used by healthcare professionals to characterise, monitor, and predict the future course of patients following a diagnosis of dementia.

## Data Availability

It is possible for external researchers to request access to the summarised (aggregated) data from this study through a formal data request process. Researchers wanting to apply to access the data from the School of Medicine, Keele University, should or contact the Principal Investigator of the study, Dr Michelle Marshall, for further information.

## Acknowledgements

The study team would also like to acknowledge the Patient and Public Involvement and Engagement Dementia Group within the School of Medicine, Keele University, for their input into the development and interpretation of results for the Course of Dementia using Medical Records (CoMed) study and also the overall Measurement of Dementia Disease Progression In Primary care (MEDDIP) programme of work, which encompasses this current study. The study team would also like to thank the Midlands Partnership NHS Foundation Trust and the project steering group for the MEDDIP study for their support.

## Funding

This work was supported by The Dunhill Medical Trust under Grant [RPGF 1711/11] as part of The MEasurement of Dementia DIsease progression in Primary care (MEDDIP) study. KPJ and CCG are also supported by matched funding awarded to the NIHR Applied Research Collaboration (West Midlands). The views and opinions expressed are those of the authors and not necessarily the views of The Dunhill Medical Trust, NHS, the NIHR or the Department of Health and Social Care.

## Declaration of interest statement

The authors have no conflicts of interest to declare.

## Author contributions

Study was derived and planned by MM, PCa, CAC-G, PCr, MF, SS, AS, KW, SW, and KPJ. MM and KPJ performed analysis. All authors contributed to interpretation of findings. MM and KPJ drafted the paper and all authors commented on subsequent draft versions and approved the final version.

## Ethical conduct of research statement

Written informed consent was obtained from patients with dementia (or personal consultee’s advice for those not able to give consent) to access and link their secondary care dementia service and primary care medical records for research purposes. Ethical approval was obtained by the UK National Research Ethics Service, Wales 7 Committee (REC reference: 18/WA/0423).

---

**Supplementary Table 1.**
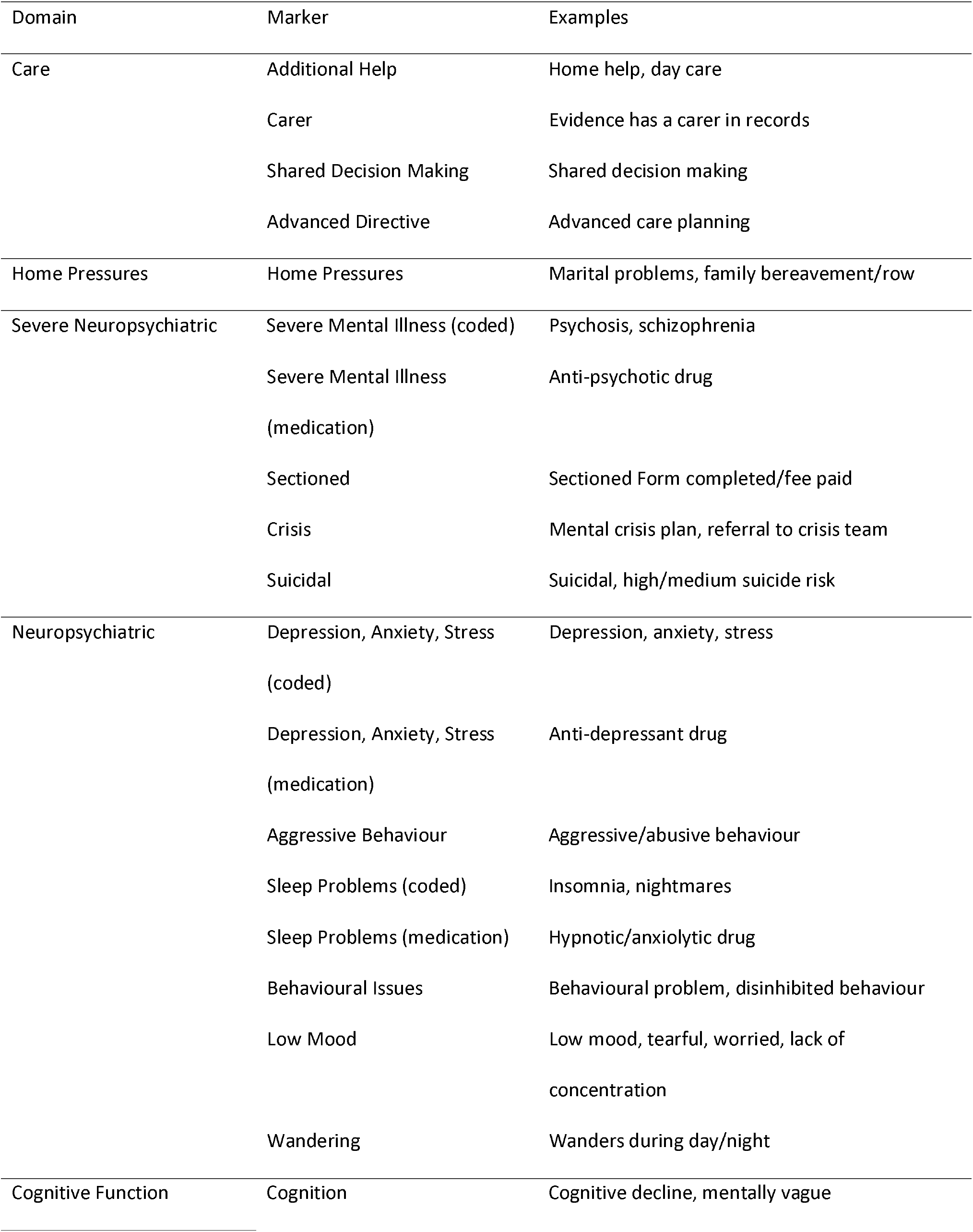
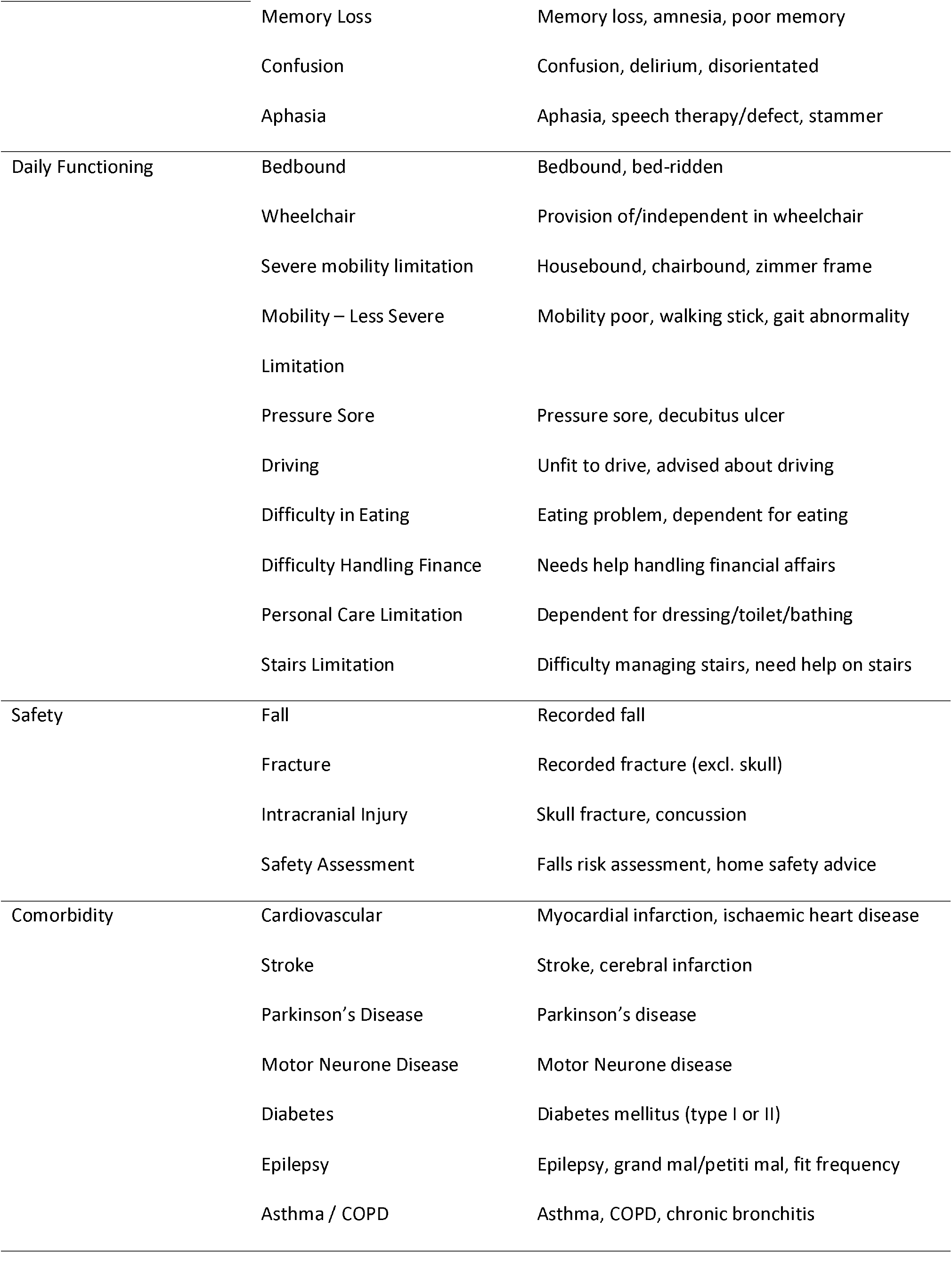
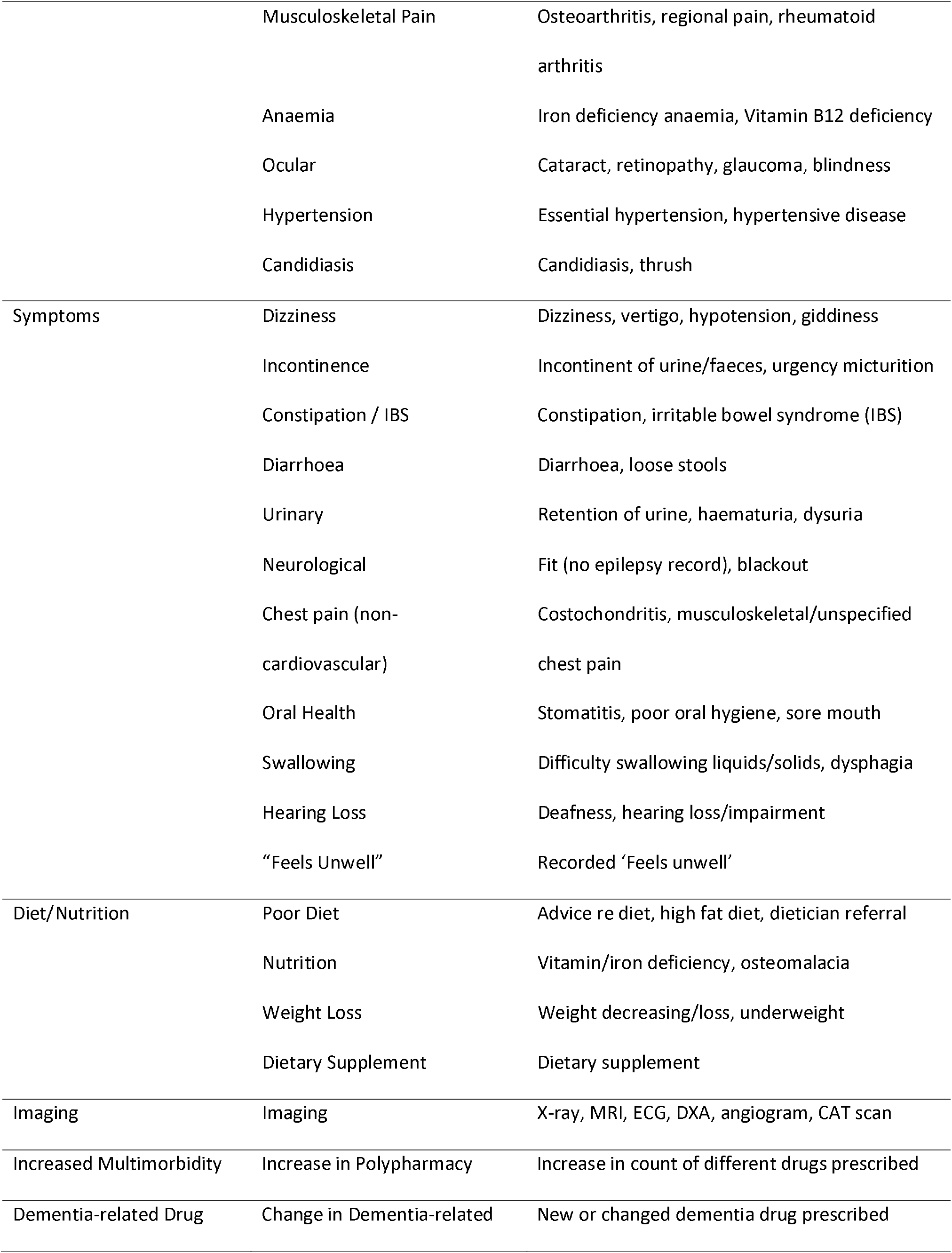
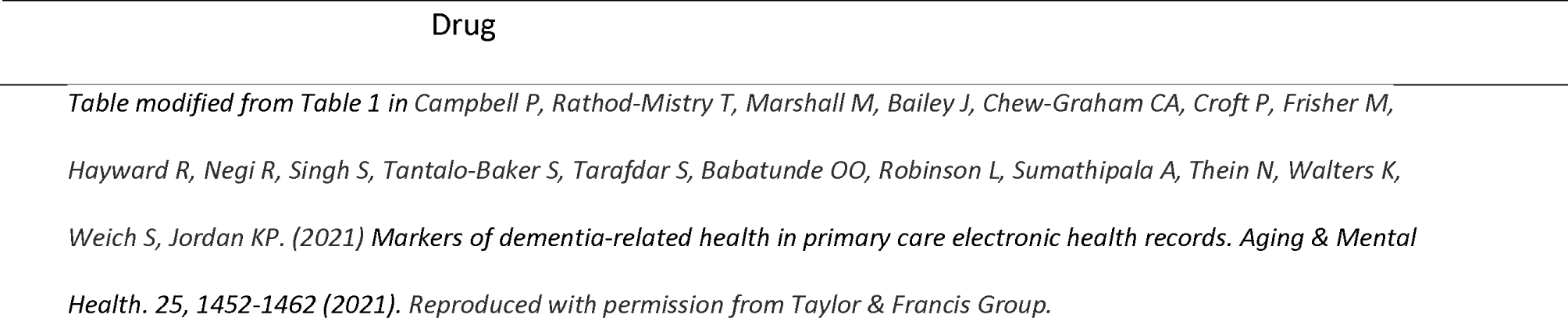
List of the markers within each domain and examples.

## Notes

### Competing Interest Statement

The authors have declared no competing interest.

### Author Declarations

Ethical approval was obtained by the UK National Research Ethics Service, Wales 7 Committee (REC reference: 18/WA/0423).

